# Development and External Validation of Multimodal Machine Learning Models to Predict High Inpatient Opioid Exposure

**DOI:** 10.64898/2026.03.31.26349842

**Authors:** Swara Kale, Devender Singh, Eeric Truumees, Matthew Geck, John Stokes

**Affiliations:** The University of Texas at Austin, 101 E 21st Street, Austin, Texas 78712, United States; Ascension Texas Spine and Scoliosis, 1004 W 32nd St #200, Austin, TX 78705, United States

## Abstract

High inpatient opioid exposure is associated with increased risk of persistent opioid use, yet early identification of high-risk patients during hospitalization remains limited. We developed and evaluated machine learning models to predict extreme opioid exposure using electronic health record data from MIMIC-IV. This retrospective cohort-based prediction modeling study included 223,452 unique first hospital admissions. The outcome was extreme opioid exposure, defined as the top decile of morphine milligram equivalents (MME) per day among opioid-exposed admissions (corresponding to ≥225 MME/day in the development cohort), representing 2.65% of all admissions.

Structured early-admission features included demographics, admission characteristics, laboratory utilization and abnormality summaries, and 24-hour procedural indicators. Discharge-note data were incorporated using ClinicalBERT embeddings and bigram features. Models were trained using an 80/10/10 split, with temporal validation performed on the most recent 10% of admissions. External validation was conducted using the MIMIC-III and eICU Collaborative Research Database cohorts. Performance was assessed using ROC-AUC and PR-AUC with 95% confidence intervals.

Among structured-only models, XGBoost achieved the best internal test performance (ROC-AUC 0.932 [0.924-0.940]; PR-AUC 0.223 [0.193-0.262]). A combined structured and notes model improved precision-recall performance (ROC-AUC 0.932 [0.920-0.943]; PR-AUC 0.276 [0.229-0.331]). Temporal validation showed similar discrimination (ROC-AUC 0.929; PR-AUC 0.223).

In external validation, performance decreased substantially. In MIMIC-III, the model achieved ROC-AUC 0.669 [0.659-0.680] and PR-AUC 0.018 [0.017-0.019], while in eICU performance was further attenuated (ROC-AUC 0.567 [0.556-0.576]; PR-AUC 0.018 [0.017-0.019]). Predicted probabilities were poorly calibrated in both external datasets, with limited correspondence between predicted and observed risk.

These findings demonstrate that while EHR-based machine learning models can achieve strong internal discrimination, their performance and calibration may degrade substantially across independent healthcare systems -- underscoring the need for dataset-specific validation and recalibration prior to clinical application.

**AUTHOR SUMMARY:** Opioid medications are commonly used in hospitals to treat pain, but some patients receive very high doses, which may increase their risk of long-term opioid use and dependence.

Identifying these high-risk patients early during hospitalization could help doctors make safer prescribing decisions and improve pain management.

In this study, we analyzed electronic health record data from over 220,000 hospital admissions to develop machine learning models that estimate which patients are likely to receive high levels of opioids. We focused on information available within the first 24 hours of admission, including patient characteristics, laboratory testing patterns, and procedures. We also examined whether information from clinical notes could improve predictions.

We found that the models performed well within the original dataset, and that combining structured data with clinical notes improved performance. The patterns identified by the models -- such as links to surgical procedures and more intensive care -- were in line with expectations. However, when the models were tested in different hospital datasets, performance declined, and predicted risks no longer matched what actually happened to patients.

These findings highlight an important challenge in applying machine learning in healthcare: models that perform well in one setting may not work reliably in others. While routinely collected hospital data may help identify patients at risk early, predictive models must be carefully tested and adapted before they can be safely used in new clinical environments.

## INTRODUCTION

Opioid use has been a critical public health concern in the United States for the past 30 years.[1, 2] Although overdose deaths have decreased in recent years, more than 68,000 individuals died from drug overdose between September 2024 and September 2025.[3, 4] Opioids may be obtained illicitly or through legitimate prescriptions. Among prescription medications, opioids remain the most frequently misused class, with more than 7.6 million prescription users transitioning to misuse annually.[5] In 2023 alone, 13,026 individuals died from prescription opioid overdose.[6]

Opioid administration during hospitalization is common, particularly following major surgical procedures and other high-intensity interventions.[7, 8] While short-term opioid therapy is often clinically necessary for acute pain management, accumulating evidence indicates that inpatient opioid exposure may increase the likelihood of persistent opioid use after discharge and elevate the risk of long-term dependence or misuse.[9–11] Additionally, substantial variability exists in inpatient opioid prescribing intensity across geographic regions, institutions, and individual prescribers, even among patients with comparable clinical presentations.[12–14] This heterogeneity suggests that factors beyond clinical necessity contribute to high-dose exposure, with some hospitalizations experiencing disproportionately intense opioid administration.

Existing efforts to mitigate opioid-related harm have largely focused on outpatient prescribing practices, prescription drug monitoring programs, and community-level interventions.[15] Comparatively less attention has been directed toward identifying patients at risk for extreme opioid exposure during hospitalization, despite the hospital setting representing a critical and potentially modifiable point of care. A predictive model capable of identifying hospitalizations at risk for unusually high opioid dosing could enable targeted stewardship and earlier clinical review.

In this study, we developed, internally validated, and externally evaluated machine learning models to predict extreme inpatient opioid exposure using electronic health record (EHR) data. Extreme exposure was defined as the top decile of morphine milligram equivalents (MME) per hospital day among opioid-exposed admissions. Our primary structured model incorporated demographic, diagnostic, procedural, laboratory, and medication-derived features available early in hospitalization to estimate the probability of unusually high opioid intensity.

We additionally evaluated a notes-only model trained on clinical documentation to determine whether unstructured text independently contains predictive signals associated with opioid intensity. Furthermore, we constructed a multimodal model integrating structured features with note-derived embeddings to assess whether combining quantitative data with clinical language improved predictive performance. Exploratory analysis of clinical text was performed to provide interpretability of text-derived signals.

Finally, we evaluated model generalizability through external validation in independent datasets. Differences in patient populations, documentation practices, and data structure across healthcare systems may substantially impact model performance. By systematically comparing structured, text-based, and multimodal approaches, we aimed to examine how model discrimination and calibration change across heterogeneous clinical settings and to assess the extent to which predictive performance translates beyond the development dataset.

## RESULTS

### Participants and Cohort Characteristics

A total of 546,028 hospital admission records were screened, of which 223,452 unique first admissions met eligibility criteria and were included in the final analytic cohort. Among these admissions, 4,627 (2.65%) met criteria for high-risk opioid exposure, defined as the top decile of daily morphine milligram equivalents (MME) among opioid-exposed admissions (corresponding to ≥225 mg/day in the development cohort). Overall, 59,441 admissions (26.5%) received any opioid exposure during hospitalization. The distribution of MME per day among opioid-exposed admissions was right-skewed (median 54.7; IQR 19.5-122.1).

Admissions were randomly assigned into training (n = 178,762; 3,730 events), validation (n = 22,345; 465 events), and test (n = 22,345; 432 events) sets, with stable outcome prevalence across splits (training: 2.09%; validation: 2.08%; test: 1.94%).

Baseline characteristics stratified by outcome status are shown in Supplementary Table 1. Compared with non-top-decile admissions, high-risk admissions were younger and had higher laboratory utilization within 24 hours. Medication administrations within 24 hours were substantially higher in the high-risk group, and surgical admissions were more frequent among high-risk cases.

### Missing Data and Feature Processing

Medication and laboratory features had moderate missingness, which was addressed using indicator variables and imputation. No residual missing values remained after preprocessing.

### Model Development and Evaluation

Structured prediction models were developed using ridge logistic regression, random forest, and extreme gradient boosting. Models were trained on the training set, tuned using the validation set, and evaluated in the test set.

Model outputs were predicted probabilities of high-risk opioid exposure, and classification thresholds were selected using the Youden index in the validation set. Primary performance metrics are summarized in Table 1. Threshold-dependent performance metrics are provided in Supplementary Table 2.

**Table 1:** Primary model performance.

| **Dataset** | **Model** | **ROC-AUC**<br>**(95% CI)** | **PR-AUC**<br>**(95% CI)** | **Brier**<br>**(95% CI)** | **Calibration**<br>**intercept** | **Calibration**<br>**slope** |
| --- | --- | --- | --- | --- | --- | --- |
| Validation | Ridge logistic | 0.892 | 0.166 | 0.0187 | — | — |
| Validation | Random forest | 0.909 | 0.201 | 0.0181 | — | — |
| Validation | XGBoost | 0.918 | 0.216 | 0.0180 | — | — |
| Validation | Notes-only | 0.865 | 0.150 | 0.0196 | — | — |
| Validation | Combined | 0.933 | 0.262 | 0.0179 | — | — |
| Test | Ridge logistic | 0.915<br>(0.903-0.925) | 0.185<br>(0.159-0.217) | 0.0172<br>(0.0158-0.0188) | 0.446 | 1.183 |
| Test | Random forest | 0.919<br>(0.909-0.928) | 0.204<br>(0.174-0.240) | 0.0169<br>(0.0155-0.0184) | 0.083 | 1.094 |
| Test | XGBoost | 0.932<br>(0.924-0.940) | 0.223<br>(0.193-0.262) | 0.0166<br>(0.0152-0.0181) | 0.123 | 1.063 |
| Test | Notes-only | 0.861<br>(0.840-0.881) | 0.168<br>(0.134-0.208) | 0.0189<br>(0.0169-0.0210) | -0.028 | 0.994 |
| Test | Combined | 0.932<br>(0.920-0.943) | 0.276<br>(0.229-0.331) | 0.0173<br>(0.0155-0.0191) | 0.036 | 1.023 |
| External<br>(MIMIC-III) | XGBoost | 0.669<br>(0.659-0.680) | 0.018<br>(0.017-0.019) | 0.0372<br>(0.0359-0.0388) | — | — |
| External<br>(eICU) | XGBoost | 0.567<br>(0.556-0.576) | 0.018<br>(0.017-0.019) | 0.0192<br>(0.0187-0.0197) | — | — |

### Model Performance: Structured-Only Models

In the independent test cohort, XGBoost achieved the best performance among structured models, with ROC-AUC 0.932 [0.924-0.940] and PR-AUC 0.223 [0.193-0.262], and demonstrated good internal calibration (slope 1.06; intercept 0.12). Random forest and ridge logistic regression showed slightly lower discrimination. Bootstrap resampling confirmed stability of performance estimates (Table 1) (Fig 1).

**Fig 1.**
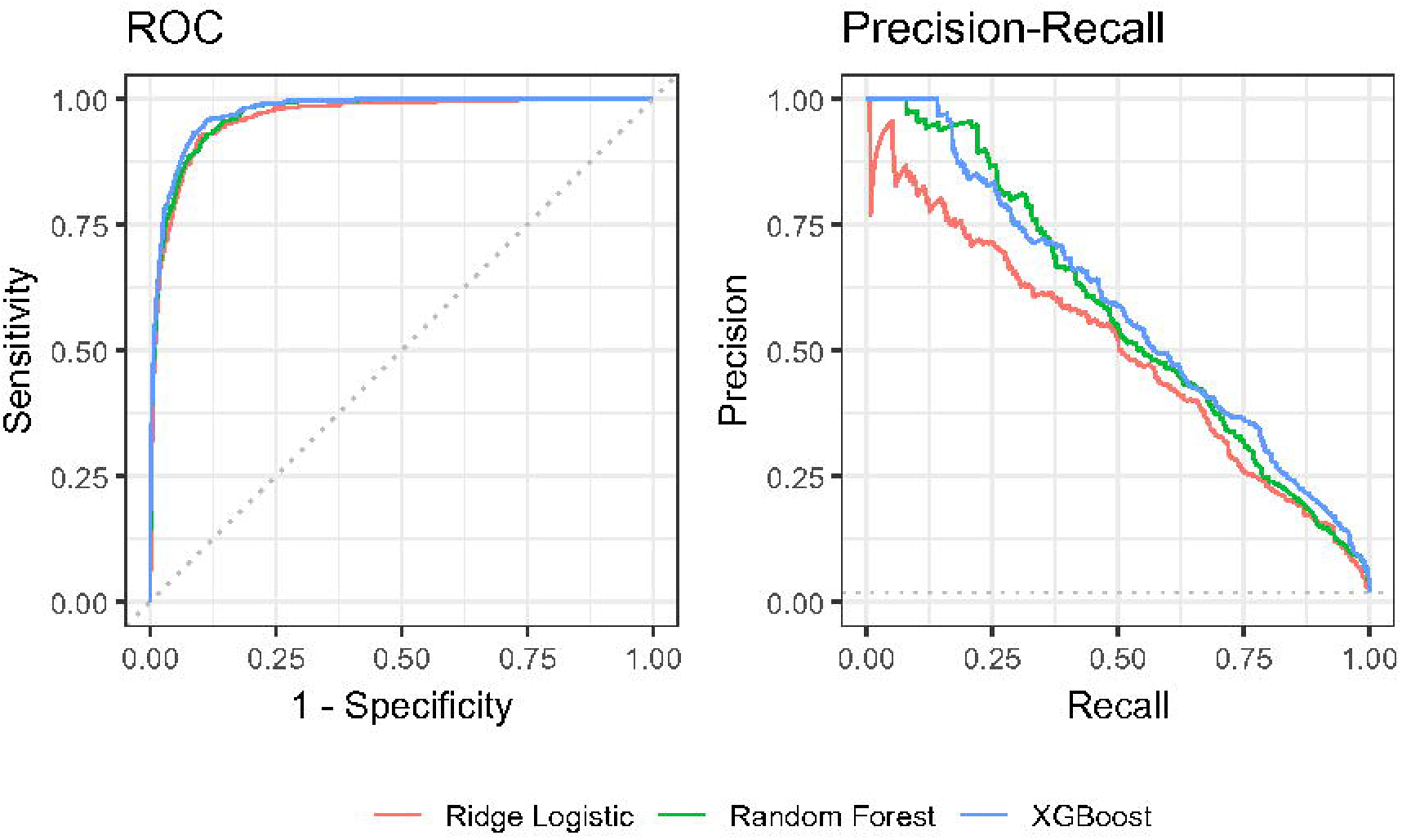
ROC and PR curves for ridge logistic regression, random forest, and XGBoost models (test set)

### Model Interpretation: SHAP Analysis

SHAP analysis of the XGBoost model indicated that early inpatient utilization features were the strongest contributors to predicted risk, particularly medication event counts, age, and surgical status (Fig 2).

**Fig 2.**
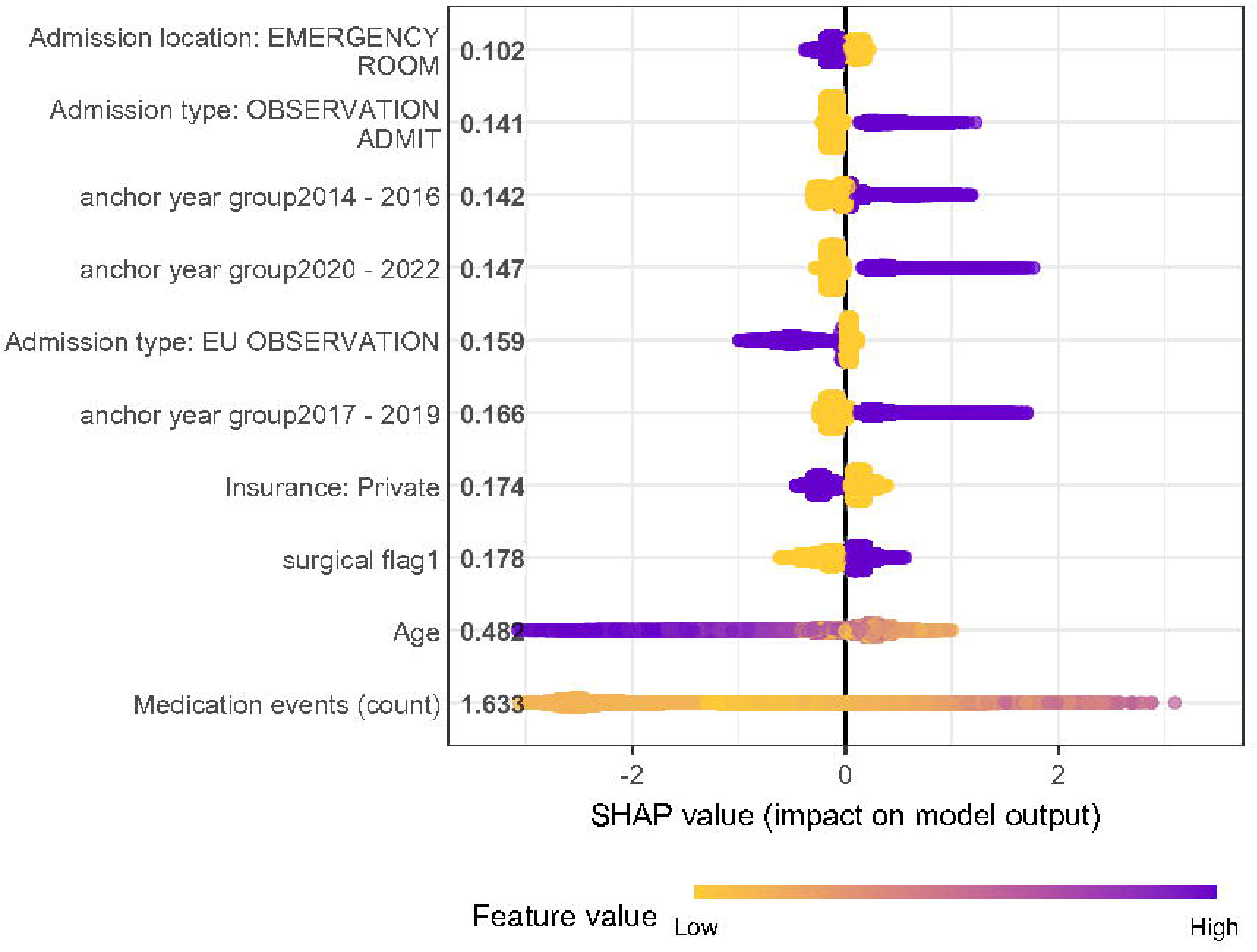
SHAP Summary plot for structured XGBoost model predicting high inpatient opioid exposure

### Notes-Only Model and Multimodal Integration

Embeddings were available for 58.4% of admissions. The notes-only model achieved ROC-AUC 0.861 and PR-AUC 0.168 in the test subset. The combined structured + notes model improved performance (ROC-AUC 0.932; PR-AUC 0.276), representing substantial enrichment over baseline prevalence (Table 1) (Fig 3). However, because discharge summaries are generated after hospitalization, these findings reflect retrospective signal capture rather than prospective prediction.

**Fig 3.**
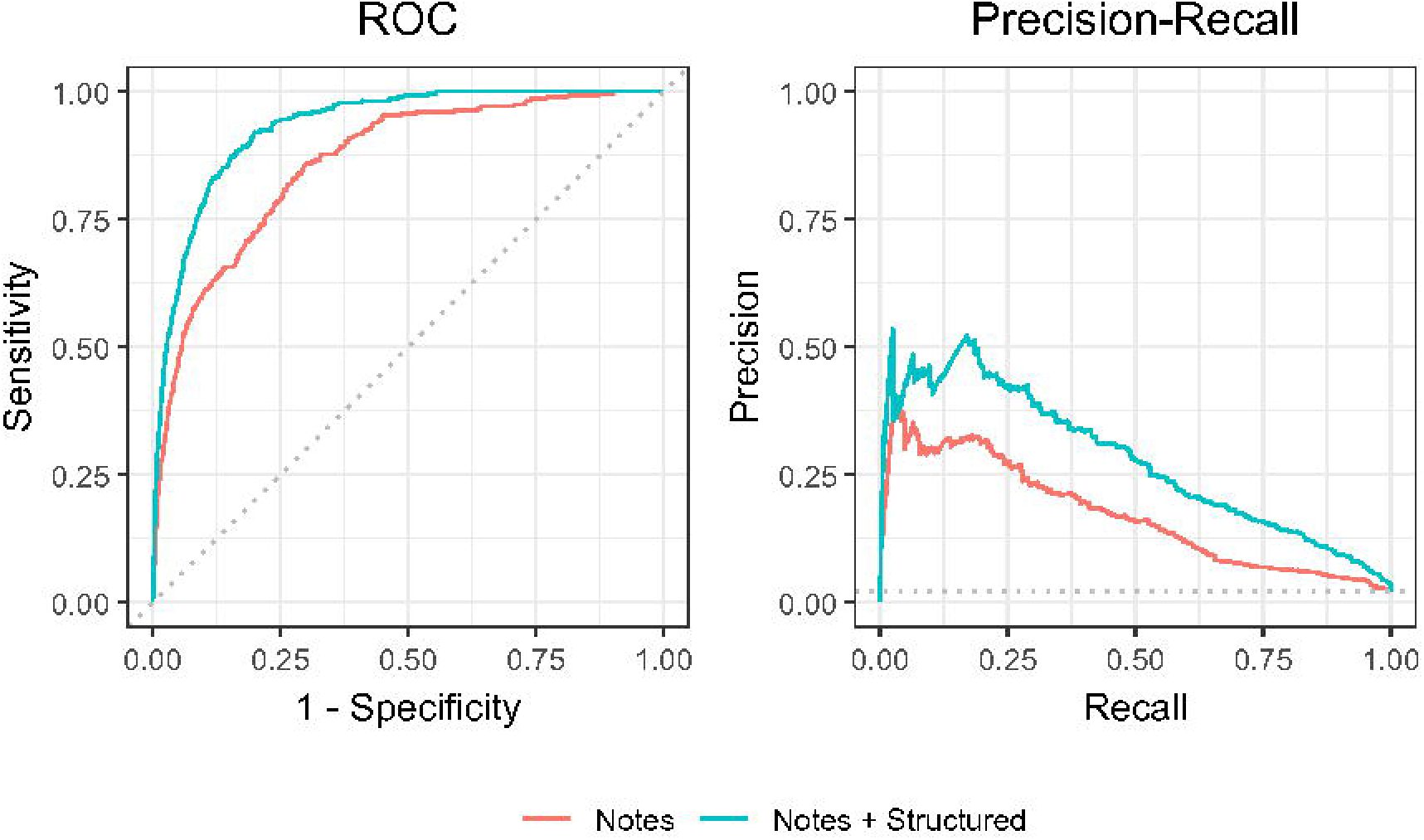
ROC and PR curves for text-based models (test set)

### Contrastive Text Analysis

Bigram analysis identified phrases enriched among high-risk admissions, including terms related to orthopedic and spine procedures (Supplementary Table 3) (Fig 4).

**Fig 4.**
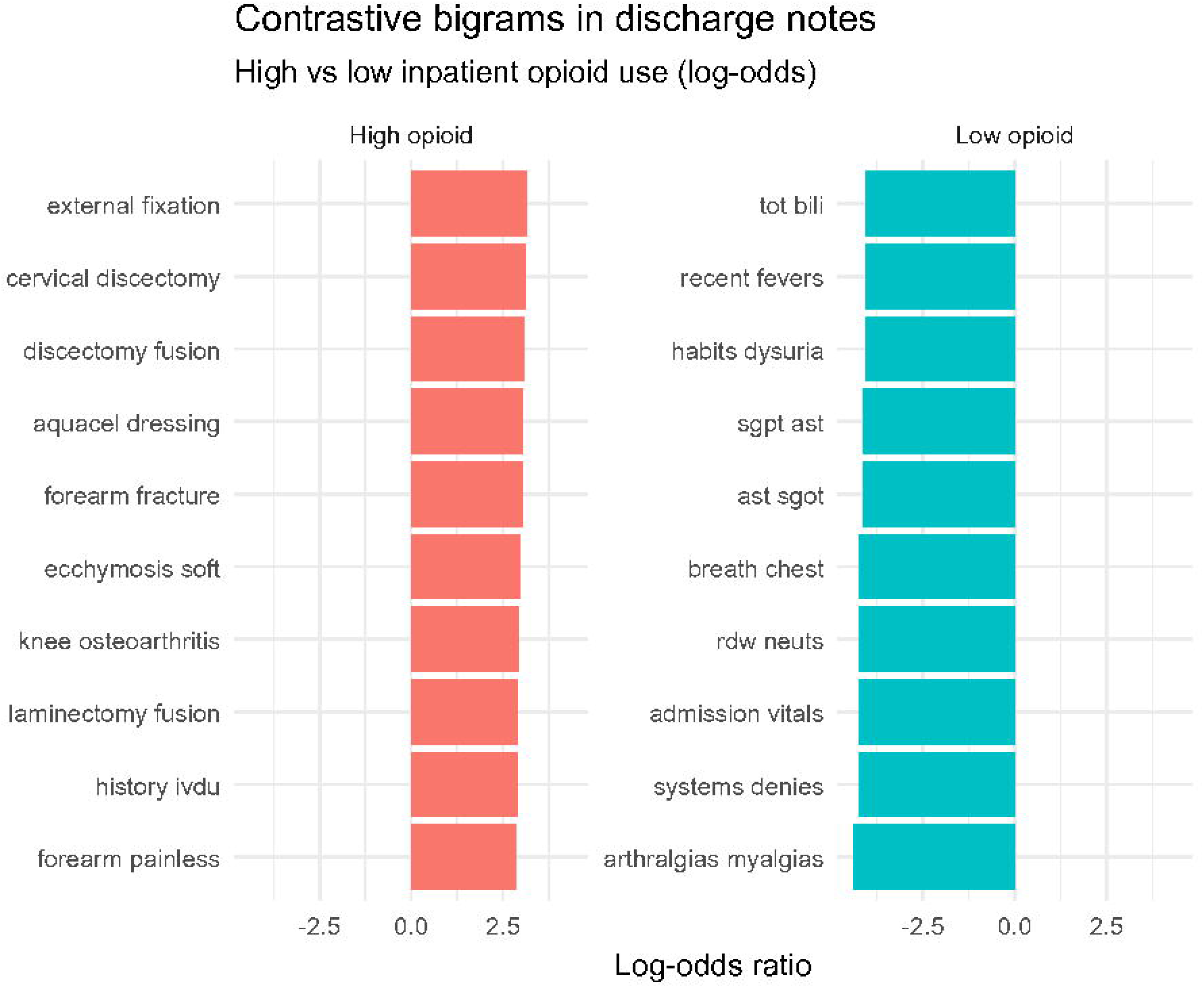
Contrastive bigrams in discharge notes associated with high versus low inpatient opioid use

### Subgroup Performance

Model performance remained stable across surgical and non-surgical admissions, with differences in PR-AUC reflecting underlying outcome prevalence. Calibration remained stable across subgroups (Supplementary Table 4) (Fig 5).

**Fig 5.**
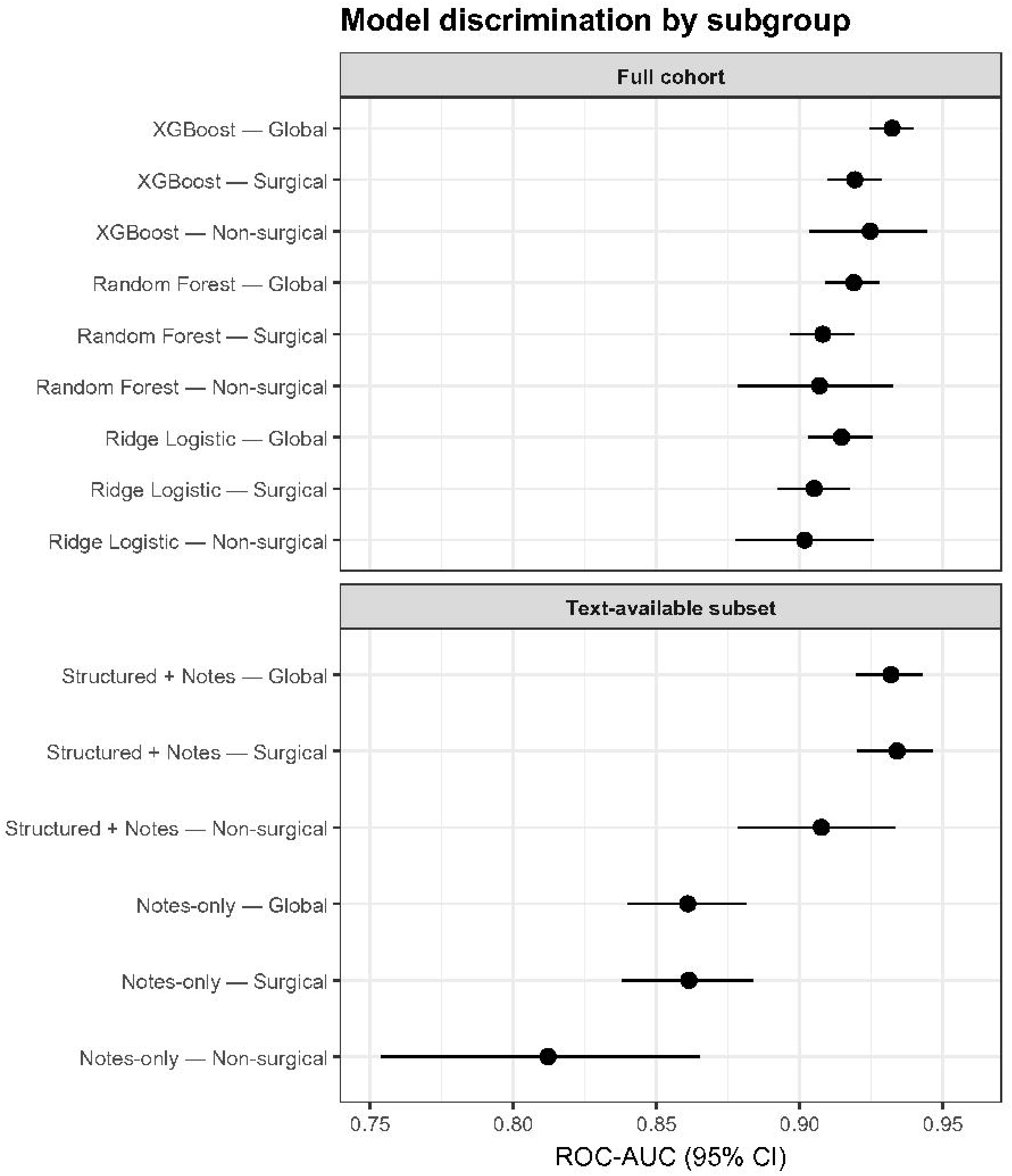
Model discrimination (ROC-AUC, 95% CI) stratified by surgical subgroup

### Temporal Validation

To assess temporal generalizability, we evaluated model performance in a temporally held-out cohort. In this cohort, XGBoost achieved ROC-AUC 0.929 and PR-AUC 0.223. Performance was similar to that observed in the primary test cohort (Table 1) (Fig 6).

**Fig 6.**
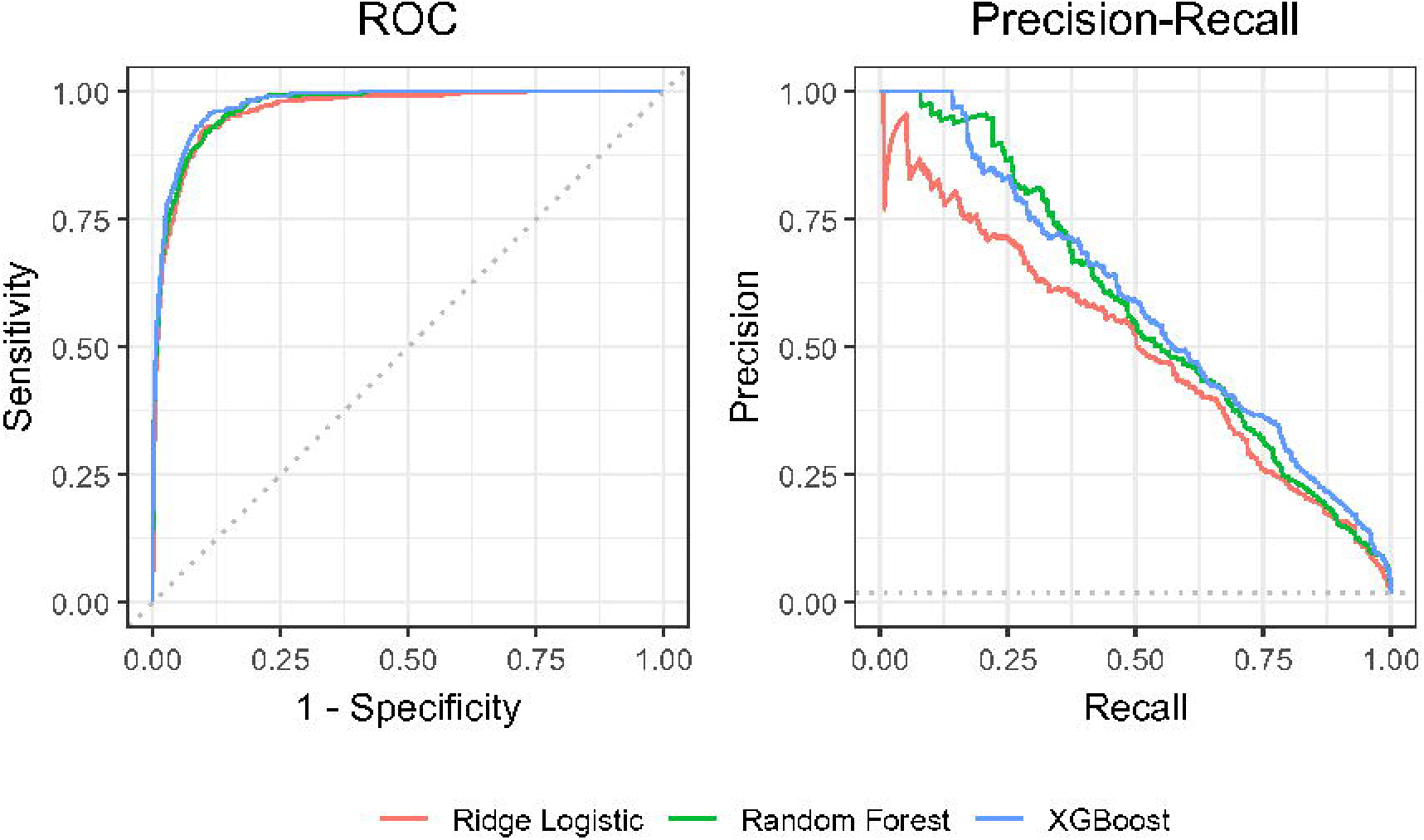
ROC and PR curves for ridge logistic regression, random forest, and XGBoost temporal validation

### External Validation

In external validation, model performance decreased substantially compared with internal and temporal evaluation. In the MIMIC-III cohort (n = 46,476; outcome prevalence 2.6%), the model achieved a ROC-AUC of 0.669 [0.659-0.680] and PR-AUC of 0.018 [0.017-0.019]. In the eICU cohort (n = 200,859; outcome prevalence 1.6%), discrimination was further attenuated, with ROC-AUC 0.567 [0.556-0.576] and PR-AUC 0.018 [0.017-0.019] approaching near-random discrimination (Table 1) (Fig 7). This represents a reduction in ROC-AUC of approximately 0.36 compared with internal test performance.

**Fig 7.**
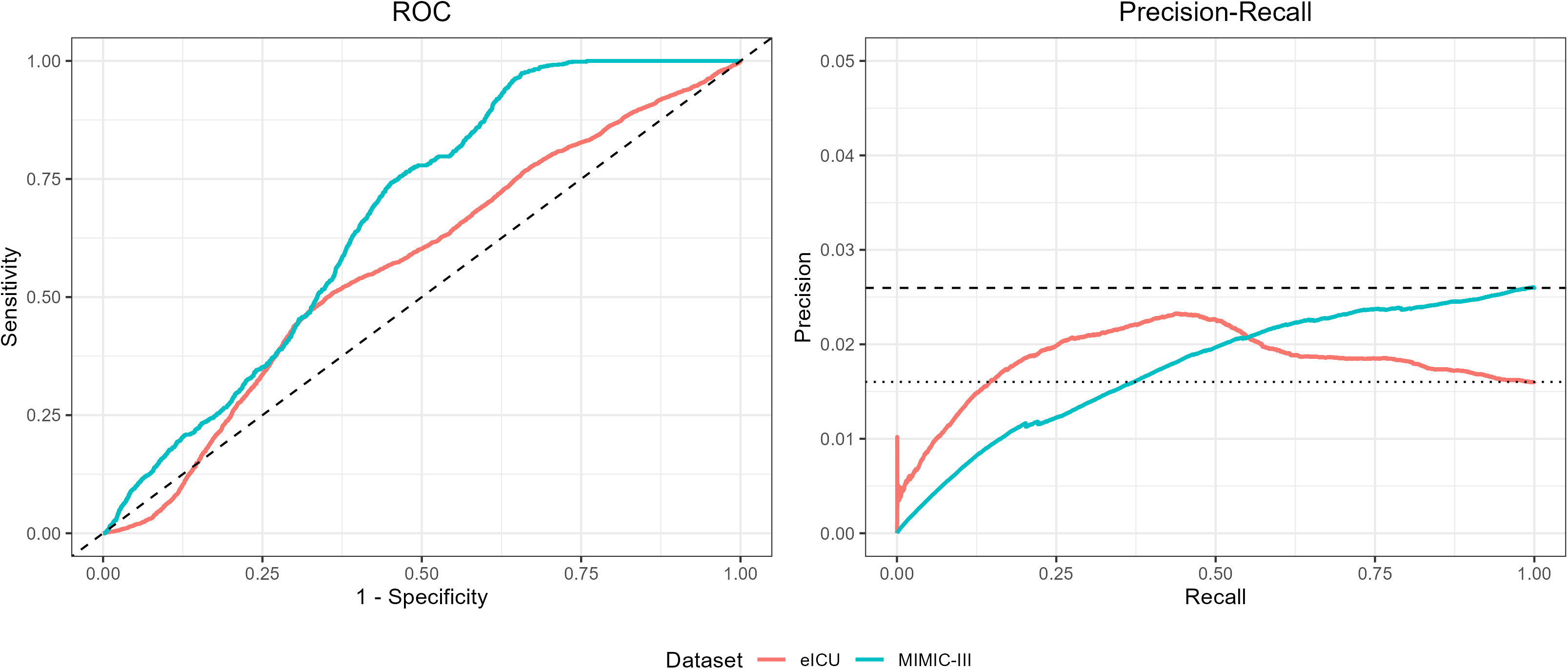
ROC and PR curves for MIMIC-III and eICU external validation XGBoost models

External calibration analysis demonstrated substantial degradation in predicted probability reliability in both external datasets. In MIMIC-III, predicted probabilities showed weak and non-monotonic association with observed outcomes, while in eICU, predicted risks exhibited minimal separation between high- and low-risk admissions. Calibration slopes were markedly reduced in both datasets, indicating severe distortion in probability scaling. These findings indicate that, although the model retained modest ranking ability, its probability estimates were not well calibrated in external settings (Fig 8).

**Fig 8.**
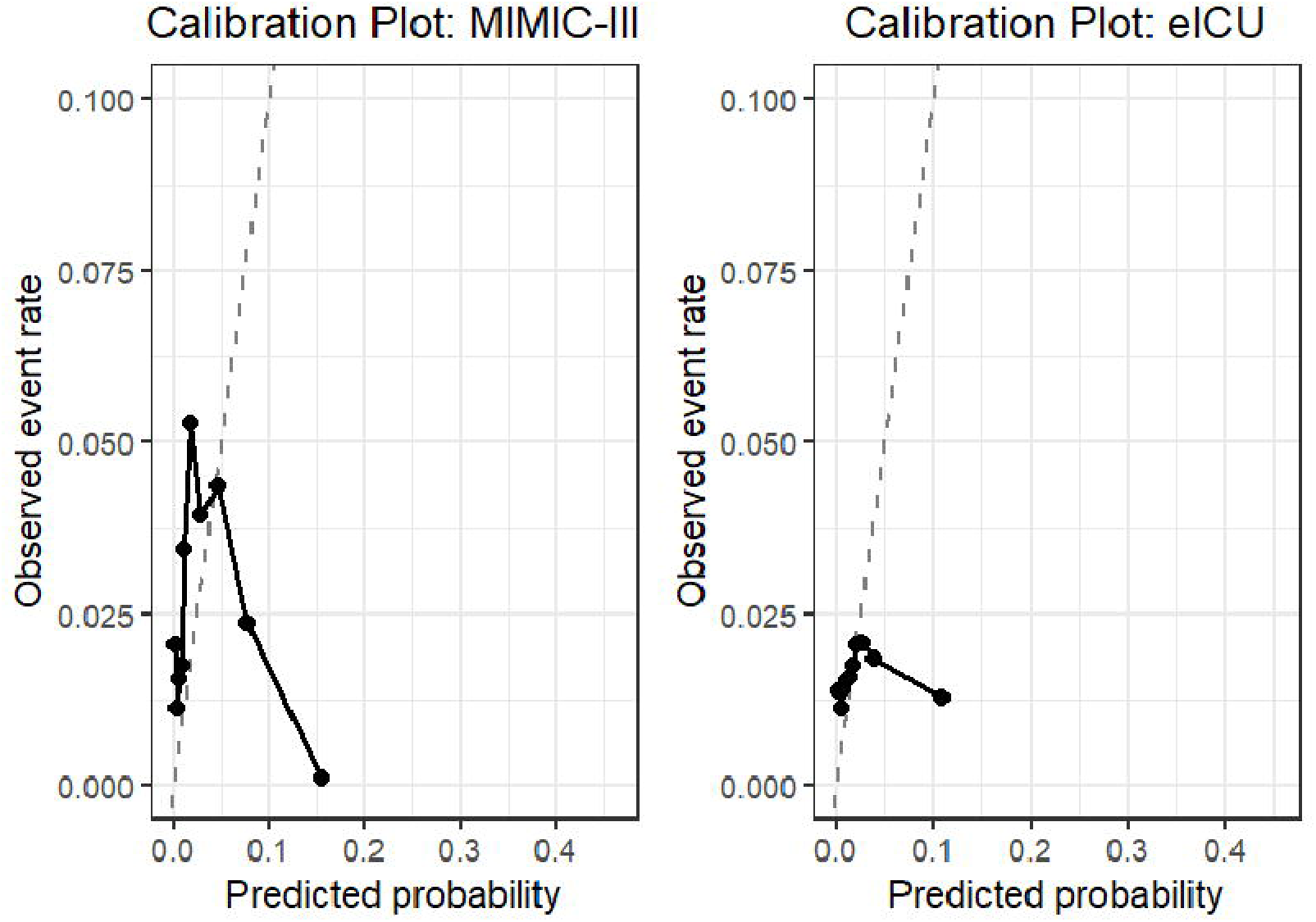
Calibration plots for MIMIC-III and eICU external validation

## DISCUSSION

In this study, we developed and validated machine learning models to identify hospital admissions at risk for extreme inpatient opioid exposure. Internal model performance demonstrated strong discrimination and suggested that routinely collected inpatient data captured broader signals of care complexity associated with high opioid exposure. However, this performance did not generalize to external datasets, where both discrimination and probability calibration declined substantially.

In clinical practice, hospitalized patients generate multimodal data streams. Structured data quantify measurable aspects of care, such as laboratory utilization, procedural burden, and medication administration, while unstructured notes capture physician interpretation, procedural narratives, and contextual detail surrounding the patient’s clinical course. These modalities represent complementary views of the same hospitalization. Our results suggest that structured utilization patterns provide a strong foundation for identifying high-intensity opioid exposure, while narrative documentation contributes additional context in specific settings, particularly surgical admissions.

Extreme opioid exposure was uncommon but clinically meaningful, occurring in a small minority of admissions. These cases were characterized by greater early laboratory utilization, higher medication event counts, and more frequent surgical procedures. Such patterns likely reflect more complex operative courses, trauma-related admissions, or high-acuity postoperative care -- contexts in which aggressive analgesia may be required.[16] Importantly, the model did not rely on opioid-specific inputs alone but instead captured broader signals of clinical intensity. This suggests that extreme dosing may be predictable from overall care trajectories rather than from pain management variables in isolation.

Subgroup analyses further illustrate how clinical context shapes predictive signals. Structured models demonstrated stable discrimination across surgical and non-surgical admissions, indicating that utilization-based features generalize across heterogeneous inpatient populations. In contrast, the notes-only model performed more strongly in surgical contexts and less consistently in non-surgical admissions. Surgical discharge summaries often contain detailed procedural narratives, implant descriptions, and postoperative plans, which may encode implicit signals of anticipated analgesic need. Medical admissions, by comparison, may contain more templated or symptom-focused language, potentially limiting the strength of narrative-based prediction. The improved performance observed with multimodal integration suggests that quantitative measures and clinician documentation capture partially distinct aspects of care complexity.

Bigram analysis of discharge notes supports the clinical plausibility of the learned narrative signals. Terms enriched among high-risk admissions were predominantly related to orthopedic, trauma, and spine procedures, such as “external fixation” and “laminectomy fusion”. These clinical scenarios are commonly associated with substantial postoperative pain and extended analgesic requirements and have been previously linked to postoperative opioid prescribing.[17] Conversely, phrases enriched among low-risk admissions reflected general review-of-systems language or laboratory descriptors.

Temporal validation demonstrated that model performance remained stable when applied to more recent admissions, suggesting that the predictive relationships identified were not limited to a specific time period. Although prescribing behaviors and stewardship efforts continue to evolve nationally, the model’s relative stability across time supports the hypothesis that extreme opioid exposure reflects persistent underlying drivers of care intensity rather than transient prescribing variation.[18]

External validation revealed a marked and clinically meaningful decline in model performance, highlighting limited transportability across independent datasets. This degradation likely reflects multiple forms of dataset shift, including differences in patient case variety, clinical practice patterns, feature availability, and data representation. For example, variation in medication documentation, laboratory reference ranges, and procedural coding required feature harmonization using proxy variables, potentially attenuating predictive signal. Additionally, differences in outcome prevalence and the use of dataset-specific percentile thresholds may have introduced label distribution shifts that further impacted performance.

Together, these results demonstrate limited transportability of the model across datasets and suggest that predictive performance is highly sensitive to differences in patient populations, feature representation, and prescribing practices. The relatively higher performance observed in MIMIC-III compared with eICU suggests partial generalizability across temporally distinct but structurally similar datasets, but poor generalization across more heterogeneous healthcare systems.

These findings suggest that a substantial portion of model performance in the development cohort may be driven by dataset-specific patterns rather than fully generalizable relationships. As a result, direct deployment of such models without recalibration may lead to inaccurate risk estimation in new clinical environments.

Methodologically, these results emphasize the importance of rigorous external validation in EHR-based machine learning studies. Models trained on a single dataset may overestimate performance due to implicit encoding of local practice patterns, documentation structures, and patient population characteristics. Future work should focus on domain adaptation, feature standardization, and recalibration strategies to improve generalizability across institutions.

From a clinical perspective, identifying admissions at risk for extreme opioid exposure has several potential applications. Because high inpatient dosing has been associated with downstream opioid use, persistent exposure, and increased healthcare utilization, early identification may represent a modifiable intervention point.[19] Operationally, a model could be deployed at the 24-hour mark following admission, once early clinical data was available.

This timepoint provides an opportunity to reassess analgesic strategies, optimize multimodal pain management, or involve specialty consultation. Given the low outcome prevalence, risk thresholds could be selected to flag only the highest-risk patients, thereby limiting alert burden while focusing attention on those most likely to experience extreme exposure. Potential future integration into clinical decision support systems could enable automated notification to care teams or pharmacy services. Importantly, predicted risk should support clinical judgment and proactive planning rather than mandate opioid restriction.

Additionally, opioid prescribing patterns vary across patient demographic characteristics.[20,21] Because predictive models may inadvertently reflect underlying practice patterns, careful evaluation across demographic subgroups is essential prior to clinical deployment. Future work should assess subgroup discrimination, calibration, and threshold performance to ensure equitable implementation and to avoid amplifying existing disparities in opioid prescribing, particularly among female patients and individuals from racial and ethnic minority backgrounds.

### Limitations

This study has several limitations. First, although external validation was performed using MIMIC-III and the eICU Collaborative Research Database, model performance declined compared with internal and temporal validation, reflecting challenges in transportability across datasets with differing patient populations, feature availability, and prescribing practices.

Second, discharge note embeddings were available for only a subset of admissions, introducing potential selection bias related to documentation availability and limiting generalizability of text-based models. Third, extreme opioid exposure was defined using a relative, dataset-specific threshold (top decile among opioid-exposed admissions), which may limit direct comparability across datasets and require recalibration in settings with different prescribing distributions. This approach also contributed to differences in outcome definition across external cohorts. Fourth, key clinical and contextual factors, including pain severity, intraoperative management, and provider-level prescribing behaviors, were not captured in the available data, potentially limiting model interpretability and performance. Finally, the low prevalence of extreme exposure limits positive predictive value, indicating that these models are best suited for risk stratification rather than definitive clinical decision-making.

## MATERIALS AND METHODS

### Data Source and Study Setting

We conducted a retrospective cohort study using MIMIC-IV for the development and validation of prediction models. MIMIC-IV is a publicly available, de-identified electronic health record (EHR) database comprising detailed clinical data from patients admitted to Beth Israel Deaconess Medical Center, a large tertiary academic medical center in Boston, Massachusetts.[22–24] The MIMIC-IV hospital module contains structured clinical data from 223,452 unique patients between 2008 and 2022, including demographic characteristics, laboratory measurements, procedures, medication administrations, and admission-level encounter data.

Free-text clinical documentation was obtained from the MIMIC-IV-Note module, which includes 331,793 de-identified discharge summaries from 145,915 patients during the same study period.[25] Discharge summaries were linked to structured admission-level data using the unique hospital admission identifier.

### Study Population

All inpatient hospital admissions recorded in the MIMIC-IV hospital module during the study period were eligible for inclusion. To avoid correlated observations from repeat hospitalizations, only the first hospital admission per patient was included in the analysis. No age-based exclusions were applied. The cohort was partitioned into training, validation, and test sets for model development, with temporal validation performed as a secondary analysis.

### Outcome Definition

The primary outcome was extreme inpatient opioid exposure within the first 24 hours of hospital admission. Opioid exposure was quantified as the total morphine milligram equivalents (MME) administered during this period, calculated using standardized equianalgesic conversion factors.[26]

Among admissions receiving any opioid within 24 hours (MME > 0), total MME values were ranked, and the 90th percentile was used to define extreme exposure. This threshold was calculated using the full cohort prior to model development and applied unchanged across all data partitions. Because this approach yields dataset-specific absolute thresholds, direct comparability across external datasets may be limited.

### Predictor Variables

Predictor domains included demographic characteristics, laboratory utilization metrics, procedure indicators, and non-opioid medication administration counts. Demographic variables included age, sex, race, marital status, insurance type, primary language, admission type, admission location, calendar year of admission, and a categorical year grouping variable.

Laboratory-related predictors included the total number of laboratory events within 24 hours, the number of laboratories with reference ranges available, counts of abnormal laboratory values (overall, low, and high), and the proportion of abnormal laboratory results. Procedure-related predictors included binary indicators for major procedural categories occurring within the first 24 hours (spine, orthopedic, neurologic, cardiac, abdominal, and vascular procedures), a composite indicator for any procedure, total procedure count, and a surgical admission flag. Medication-related predictors explicitly excluded opioid administrations and opioid-derived features. Variables directly representing opioid dose or opioid administration counts were excluded from predictors. In secondary analyses, free-text discharge summaries were used to construct text-based predictors. As discharge summaries are authored after hospitalization and may contain post-exposure information, text-based models were evaluated as retrospective comparators rather than real-time early prediction tools.

### Data Preprocessing and Feature Engineering

All preprocessing steps were performed after partitioning the dataset into training, validation, and test sets. Structured predictors were derived from data available within the first 24 hours of admission and aggregated at the admission level.

Laboratory-related variables were treated as structurally zero when no laboratory values were recorded during the 24-hour window. Indicator variables were created to denote admissions with no laboratory measurements and no laboratory results with reference ranges.

For remaining numeric predictors, missing values were imputed using median imputation. Median imputation values were derived from the training dataset and applied unchanged to validation and test sets.

Categorical predictors were encoded as factor variables. Missing or empty categorical values were assigned to a separate “Unknown” category. Factor levels were defined using the training dataset and applied consistently to validation and test datasets to ensure alignment across partitions.

All analyses were conducted in R (version 4.4.2) and Python (version 3.12.10) using ranger, glmnet, xgboost, and HuggingFace Transformers libraries.[28–32]

### Discharge Notes Preprocessing

Text representations were generated using a pretrained ClinicalBERT model, a domain-adapted transformer model trained on clinical text.[33] Embeddings for validation and test sets were generated without exposure to outcome labels and without updating model weights.

A front-back truncation strategy was applied. For each note, two tokenized segments were generated: 1) the first 256 tokens of the note (front segment) and 2) the last 256 tokens of the note (back segment).

Each segment was independently processed using ClinicalBERT, and the embedding corresponding to the [CLS] token was extracted as a 768-dimensional representation. Front and back embeddings were concatenated to produce a 1,536-dimensional vector for each admission-level note. Resulting embeddings were merged with structured admission-level predictors for downstream modeling.

### Data Partitioning

The cohort was randomly partitioned into training (80%), validation (10%), and test (10%) sets using stratified sampling to preserve outcome prevalence across partitions. The validation set was used for hyperparameter tuning and model selection. The test set was held out and used exclusively for final performance evaluation. The random stratified split served as the primary evaluation framework, with temporal validation conducted as a secondary analysis to assess robustness to distributional shift.

### Models

Three model classes were evaluated for structured predictors: random forest, penalized logistic regression, and gradient boosting. For text-only models, admission-level ClinicalBERT embeddings were used as predictors in a ridge-regularized logistic regression model. For combined models, structured predictors and text embeddings were concatenated and modeled using ridge-regularized logistic regression to stabilize estimation in the presence of high-dimensional features and to reduce overfitting. The XGBoost model was selected as the primary model for external validation based on internal performance.

### Hyperparameter Tuning

Random forest models were trained using the ranger implementation with 500 trees. Class imbalance was addressed using inverse-prevalence class weights computed from the training data. The regularization parameter (λ) for ridge-regularized logistic regression models was selected using 5-fold cross-validation. For gradient boosting models, hyperparameters were tuned using the validation set, including learning rate, tree depth, subsampling, and column sampling, with early stopping based on validation ROC-AUC. For text-only and combined models, ridge regularization was selected via cross-validation on the training set.

### Model Evaluation

Model performance was evaluated on the held-out test set. Discrimination was assessed using the area under the receiver operating characteristic curve (ROC-AUC) and the area under the precision-recall curve (PR-AUC), the latter included given the relatively low prevalence of the outcome. 95% confidence intervals were estimated using nonparametric bootstrapping with 2,000 resamples of the test set.

Overall prediction error was quantified using the Brier score. Calibration in external datasets was assessed using calibration curves and calibration slope and intercept.

### Model Interpretation

To characterize feature contributions, SHAP values were computed for the XGBoost model to quantify the relative importance and directionality of individual predictors in relation to high opioid exposure.[34]

### Subgroup Analysis

To evaluate performance across clinically distinct admission types, analyses were conducted for global, surgical, and non-surgical cohorts. A binary surgical admission flag was derived using International Classification of Diseases (ICD) procedure codes recorded for each hospital admission.[35, 36] Using the surgical flag, separate models were trained and evaluated within the surgical-only and non-surgical-only cohorts. The original global exposure threshold was retained for subgroup analyses. The same modeling pipeline, hyperparameter selection strategy, and evaluation framework described for the global models were applied to each subgroup. For each subgroup, discrimination, calibration, and clinical utility metrics were reported using the same procedures applied to the full cohort.

### Temporal Validation

To evaluate robustness to temporal dataset shift, a time-based split was performed using admission timestamp, with the most recent 10% of admissions reserved as a temporally held-out test cohort. Imputation parameters, categorical level definitions, and any high-cardinality category handling were derived from the temporal training set and applied unchanged to the temporal validation and test sets to prevent information leakage.

Random forest, ridge-regularized logistic regression, and XGBoost models were retrained using the temporal training set. Hyperparameter selection and early stopping procedures mirrored those used in the primary random split analysis. Final performance was evaluated on the temporal test set (latest 10% of admissions) using matching discrimination and calibration metrics.

### Contrastive Text Analysis

Exploratory contrastive text analysis was performed on discharge summaries to identify language patterns associated with high versus low opioid exposure. Discharge notes were tokenized and converted into bigrams. Tokens were restricted to alphabetic words 3-25 characters in length, with standard English stopwords and a curated set of clinical stopwords removed. Repeated-word bigrams were excluded, as were bigrams occurring fewer than 100 times overall. For each retained bigram, a log-odds ratio with Dirichlet smoothing was computed to quantify enrichment in high- versus low-opioid groups.

### External Validation

To evaluate model generalizability across independent datasets, external validation was performed using the MIMIC-III and eICU Collaborative Research Database cohorts.[37–40]

For MIMIC-III, an ICU-based cohort was constructed using first ICU stays per patient. The final cohort included 46,476 ICU admissions, with an outcome prevalence of 2.6% (n = 1,207). For eICU, we utilized the multicenter ICU cohort comprising 200,859 admissions across diverse hospital systems, with an outcome prevalence of 1.6% (n = 3,218).

Predictor variables for external validation were derived using data available within the first 24 hours of ICU admission, consistent with the feature definitions used in the MIMIC-IV development cohort. Due to differences in database structure and variable availability, predictors were harmonized across datasets using analogous data elements where direct equivalents were unavailable. Laboratory features were derived from 24-hour lab records as counts of measurements and abnormal values, with abnormality defined using available flags in MIMIC-III and approximated using total lab counts in eICU. Medication features were constructed using administration records and counts of opioid-related medication events within the first 24 hours were used as proxies for treatment intensity. Procedure-related features were derived from procedure and treatment tables, including counts of procedures and binary indicators of any procedure within 24 hours. A surgical admission flag was defined based on the presence of procedure codes in MIMIC-III and approximated in eICU using admission diagnosis text.

The primary outcome, extreme opioid exposure within 24 hours, was defined independently within each dataset using the same percentile-based approach as in the development cohort. Among admissions receiving any opioid within the first 24 hours, the top decile of total MME was used to define extreme exposure, ensuring consistency in relative outcome definition, while allowing absolute exposure thresholds to vary across datasets.

The final XGBoost model trained on the MIMIC-IV development cohort was applied directly to external datasets without retraining. No model recalibration was performed prior to external evaluation. All preprocessing steps, including median imputation values, categorical level definitions, and feature encoding schemes, were derived from the MIMIC-IV training data and applied unchanged to MIMIC-III and eICU to prevent information leakage.

Model performance was evaluated using ROC-AUC and PR-AUC. Confidence intervals were estimated using nonparametric bootstrapping (B = 500). External validation results were compared with internal and temporal validation to assess generalizability across datasets.

## Data Availability

The data used in this study is available from the MIMIC-IV, MIMIC-III, and eICU databases, which are hosted on PhysioNet (https://physionet.org/).

## Acknowledgments

The authors acknowledge the PhysioNet team and the investigators who contributed to the MIMIC-IV, MIMIC-III, and eICU databases.[22–25][37–40] Authors completed the required data use training and certification for access.

## Competing Interests

The authors declare that they have no known competing financial interests or personal relationships that could have appeared to influence the work reported in this paper.

## Funding Disclosures

The authors declare that they have no funding disclosures to report.

## Notes

### Competing Interest Statement

The authors have declared no competing interest.

### Funding Statement

The author(s) received no specific funding for this work.

### Summary of Updates

This revised manuscript incorporates several substantive updates to address prior feedback and strengthen the overall contribution. Most importantly, we added external validation using two independent cohorts, MIMIC III and eICU, to evaluate generalizability beyond the original development dataset. These cohorts differ in patient population and care setting, allowing assessment of model transportability across ICU based and multicenter environments. External validation results are now presented with discrimination and calibration metrics, including ROC AUC, PR AUC, Brier score, calibration slope, and intercept, along with corresponding confidence intervals.

