## Supplemental Tables 1-4 for "Development and External Validation of Multimodal Machine Learning Models to Predict High Inpatient Opioid Exposure"

S1: Baseline characteristics of full cohort[1]

| Feature | Non-Top Decile | Top Decile of All Opioid-Prescribed Patients | p-value |
|---|---|---|---|
| Age (years) | 57 (39-72) | 52 (38-62) | <0.001 |
| Anchor year | 2,151 (2,131-2,171) | 2,151 (2,131-2,170) | 0.018 |
| Laboratory measurements (count-0–24h) | 33 (23-55) | 36 (24-64) | <0.001 |
| Labs with reference ranges (count-0–24h) | 31 (21-51) | 32 (22-56) | <0.001 |
| Abnormal labs (count-0–24h) | 9 (4-17) | 10 (5-20) | <0.001 |
| Low abnormal labs (count-0–24h) | 4 (1-8) | 5 (3-10) | <0.001 |
| High abnormal labs (count-0–24h) | 5 (2-10) | 5 (2-11) | <0.001 |
| Medication administrations (count-0–24h) | 30 (18-44) | 49 (36-63) | <0.001 |
| Spine procedure within 24h | 5,507 (2.5%) | 284 (4.8%) | <0.001 |
| Orthopedic procedure within 24h | 4,050 (1.9%) | 508 (8.6%) | <0.001 |
| Neurosurgical procedure within 24h | 936 (0.4%) | 13 (0.2%) | 0.018 |
| Cardiac procedure within 24h | 5,005 (2.3%) | 69 (1.2%) | <0.001 |
| Abdominal procedure within 24h | 1,231 (0.6%) | 5 (<0.1%) | <0.001 |
| Vascular procedure within 24h | 2,295 (1.1%) | 3 (<0.1%) | <0.001 |

| | | | |
|---|---|---|---|
| Any procedure within 24h | 40,633 (19%) | 1,359 (23%) | <0.001 |
| Opioid dose (MME-0–24h) | 0 (0-0) | 231 (128-344) | <0.001 |
| Opioid administrations (count-0–24h) | 0.0 (0.0-0.0) | 13.0 (8.0-20.0) | <0.001 |
| Admission type | | | <0.001 |
| AMBULATORY OBSERVATION | 2,817 (1.3%) | 30 (0.5%) | |
| DIRECT EMER. | 4,516 (2.1%) | 110 (1.9%) | |
| DIRECT OBSERVATION | 7,944 (3.7%) | 372 (6.3%) | |
| ELECTIVE | 4,078 (1.9%) | 66 (1.1%) | |
| EU OBSERVATION | 52,939 (24%) | 570 (9.6%) | |
| EW EMER. | 72,611 (33%) | 1,285 (22%) | |
| OBSERVATION ADMIT | 25,316 (12%) | 1,878 (32%) | |
| SURGICAL SAME DAY ADMISSION | 19,585 (9.0%) | 1,065 (18%) | |
| URGENT | 27,724 (13%) | 546 (9.2%) | |
| Admission location | | | <0.001 |
| AMBULATORY SURGERY TRANSFER | 83 (<0.1%) | 9 (0.2%) | |
| CLINIC REFERRAL | 2,858 (1.3%) | 78 (1.3%) | |
| EMERGENCY ROOM | 105,101 (48%) | 1,181 (20%) | |

| | | | |
|---|---|---|---|
| INFORMATION NOT AVAILABLE | 217 (<0.1%) | 25 (0.4%) | |
| INTERNAL TRANSFER TO OR FROM PSYCH | 79 (<0.1%) | 0 (0%) | |
| PACU | 2,269 (1.0%) | 49 (0.8%) | |
| PHYSICIAN REFERRAL | 58,438 (27%) | 2,298 (39%) | |
| PROCEDURE SITE | 3,631 (1.7%) | 30 (0.5%) | |
| TRANSFER FROM HOSPITAL | 31,843 (15%) | 1,762 (30%) | |
| TRANSFER FROM SKILLED NURSING FACILITY | 946 (0.4%) | 25 (0.4%) | |
| WALK-IN/SELF REFERRAL | 12,064 (5.5%) | 465 (7.9%) | |
| Insurance | | | <0.001 |
| Medicaid | 35,838 (17%) | 1,748 (30%) | |
| Medicare | 80,179 (38%) | 1,569 (27%) | |
| No charge | 215 (0.1%) | 0 (0%) | |
| Other | 6,869 (3.3%) | 272 (4.7%) | |
| Private | 87,446 (42%) | 2,210 (38%) | |
| Primary language | | | <0.001 |
| American Sign Language | 152 (<0.1%) | 0 (0%) | |
| Amharic | 104 (<0.1%) | 0 (0%) | |
| Arabic | 294 (0.1%) | 9 (0.2%) | |

| | | |
|---|---|---|
| Armenian | 45 (<0.1%) | 0 (0%) |
| Bengali | 66 (<0.1%) | 1 (<0.1%) |
| Chinese | 3,348 (1.5%) | 21 (0.4%) |
| English | 197,073 (91%) | 5,640 (95%) |
| French | 81 (<0.1%) | 1 (<0.1%) |
| Haitian | 991 (0.5%) | 12 (0.2%) |
| Hindi | 95 (<0.1%) | 0 (0%) |
| Italian | 301 (0.1%) | 1 (<0.1%) |
| Japanese | 92 (<0.1%) | 0 (0%) |
| Kabuverdianu | 1,736 (0.8%) | 14 (0.2%) |
| Khmer | 102 (<0.1%) | 1 (<0.1%) |
| Korean | 160 (<0.1%) | 0 (0%) |
| Modern Greek (1453-) | 274 (0.1%) | 3 (<0.1%) |
| Other | 601 (0.3%) | 7 (0.1%) |
| Persian | 164 (<0.1%) | 2 (<0.1%) |
| Polish | 123 (<0.1%) | 0 (0%) |
| Portuguese | 1,400 (0.6%) | 52 (0.9%) |
| Russian | 2,411 (1.1%) | 4 (<0.1%) |
| Somali | 36 (<0.1%) | 0 (0%) |
| Spanish | 6,639 (3.1%) | 144 (2.4%) |
| Thai | 121 (<0.1%) | 3 (<0.1%) |

| | | | |
|---|---|---|---|
| Vietnamese | 493 (0.2%) | 1 (<0.1%) | |
| Marital status | | | <0.001 |
| DIVORCED | 13,258 (6.4%) | 522 (9.7%) | |
| MARRIED | 93,777 (45%) | 2,103 (39%) | |
| SINGLE | 80,494 (39%) | 2,541 (47%) | |
| WIDOWED | 18,938 (9.2%) | 225 (4.2%) | |
| Race | | | <0.001 |
| AMERICAN INDIAN/ALASKA NATIVE | 460 (0.2%) | 10 (0.2%) | |
| ASIAN | 4,457 (2.0%) | 33 (0.6%) | |
| ASIAN - ASIAN INDIAN | 750 (0.3%) | 12 (0.2%) | |
| ASIAN - CHINESE | 3,188 (1.5%) | 24 (0.4%) | |
| ASIAN - KOREAN | 316 (0.1%) | 3 (<0.1%) | |
| ASIAN - SOUTH EAST ASIAN | 774 (0.4%) | 4 (<0.1%) | |
| BLACK/AFRICAN | 1,398 (0.6%) | 31 (0.5%) | |
| BLACK/AFRICAN AMERICAN | 22,908 (11%) | 453 (7.6%) | |
| BLACK/CAPE VERDEAN | 2,138 (1.0%) | 26 (0.4%) | |
| BLACK/CARIBBEAN ISLAND | 1,388 (0.6%) | 24 (0.4%) | |
| HISPANIC OR LATINO | 4,084 (1.9%) | 1 (<0.1%) | |

| | | |
|---|---|---|
| HISPANIC/LATINO - CENTRAL AMERICAN | 191 (<0.1%) | 6 (0.1%) |
| HISPANIC/LATINO - COLUMBIAN | 351 (0.2%) | 6 (0.1%) |
| HISPANIC/LATINO - CUBAN | 186 (0.1%) | 6 (0.1%) |
| HISPANIC/LATINO - DOMINICAN | 2,164 (1.0%) | 56 (0.9%) |
| HISPANIC/LATINO - GUATEMALAN | 652 (0.3%) | 15 (0.3%) |
| HISPANIC/LATINO - HONDURAN | 235 (0.1%) | 3 (<0.1%) |
| HISPANIC/LATINO - MEXICAN | 407 (0.2%) | 15 (0.3%) |
| HISPANIC/LATINO - PUERTO RICAN | 3,014 (1.4%) | 116 (2.0%) |
| HISPANIC/LATINO - SALVADORAN | 484 (0.2%) | 11 (0.2%) |
| MULTIPLE RACE/ETHNICITY | 327 (0.2%) | 0 (0%) |
| NATIVE HAWAIIAN OR OTHER PACIFIC ISLANDER | 248 (0.1%) | 9 (0.2%) |
| OTHER | 9,216 (4.2%) | 248 (4.2%) |
| PATIENT DECLINED TO ANSWER | 1,437 (0.7%) | 31 (0.5%) |
| PORTUGUESE | 603 (0.3%) | 26 (0.4%) |
| SOUTH AMERICAN | 311 (0.1%) | 7 (0.1%) |
| UNABLE TO OBTAIN | 2,761 (1.3%) | 99 (1.7%) |
| UNKNOWN | 10,945 (5.0%) | 528 (8.9%) |
| WHITE | 134,472 (62%) | 3,853 (65%) |
| WHITE - BRAZILIAN | 655 (0.3%) | 21 (0.4%) |

| | | | |
|---|---|---|---|
| WHITE - EASTERN EUROPEAN | 596 (0.3%) | 22 (0.4%) | |
| WHITE - OTHER EUROPEAN | 4,719 (2.2%) | 209 (3.5%) | |
| WHITE - RUSSIAN | 1,695 (0.8%) | 14 (0.2%) | |
| Sex | | | <0.001 |
| F | 115,111 (53%) | 2,625 (44%) | |
| M | 102,419 (47%) | 3,297 (56%) | |
| Anchor year group | | | <0.001 |
| 2008 - 2010 | 66,730 (31%) | 180 (3.0%) | |
| 2011 - 2013 | 46,870 (22%) | 179 (3.0%) | |
| 2014 - 2016 | 41,391 (19%) | 1,397 (24%) | |
| 2017 - 2019 | 35,628 (16%) | 2,400 (41%) | |
| 2020 - 2022 | 26,911 (12%) | 1,766 (30%) | |
| Surgical admission | | | <0.001 |
| 0 | 107,513 (49%) | 1,749 (30%) | |
| 1 | 110,017 (51%) | 4,173 (70%) | |

[1]Continuous variables are reported as median (interquartile range)

### S2: Secondary model performance (test set)

| Model | Sensitivity (95% CI) | Specificity (95% CI) | PPV (95% CI) | NPV (95% CI) |
|---|---|---|---|---|
| Ridge logistic | 0.876 (0.836-0.911) | 0.822 (0.802-0.857) | 0.089 (0.078-0.106) | 0.997 (0.996-0.998) |
| Random forest | 0.855 (0.830-0.958) | 0.838 (0.723-0.859) | 0.095 (0.063-0.109) | 0.997 (0.996-0.999) |
| XGBoost | 0.906 (0.880-0.946) | 0.828 (0.790-0.845) | 0.094 (0.079-0.108) | 0.998 (0.997-0.999) |
| Notes-only | 0.859 (0.750-0.897) | 0.701 (0.672-0.812) | 0.058 (0.052-0.079) | 0.996 (0.993-0.997) |
| Combined | 0.920 (0.826-0.948) | 0.801 (0.792-0.889) | 0.091 (0.082-0.146) | 0.998 (0.996-0.999) |

### S3: Top 30 bigrams associated with high and low opioid exposure[1]

| Direction | Bigram | Admissions (low exposure) containing bigram | Admissions (high exposure) containing bigram | Total | Log odds[1] |
|---|---|---|---|---|---|
| High exposure | external fixation | 123 | 37 | 160 | 3.156 |
| High exposure | cervical discectomy | 82 | 23 | 105 | 3.098 |
| High exposure | discectomy fusion | 90 | 25 | 115 | 3.086 |
| High exposure | aquacel dressing | 95 | 25 | 120 | 3.033 |
| High exposure | forearm fracture | 84 | 22 | 106 | 3.032 |
| High exposure | ecchymosis soft | 186 | 47 | 233 | 2.979 |
| High exposure | knee osteoarthritis | 241 | 57 | 298 | 2.910 |
| High exposure | laminectomy fusion | 246 | 58 | 304 | 2.907 |

| | | | | | |
|---|---|---|---|---|---|
| High exposure | history ivdu | 127 | 29 | 156 | 2.888 |
| High exposure | forearm painless | 136 | 30 | 166 | 2.853 |
| High exposure | interbody fusion | 116 | 25 | 141 | 2.835 |
| High exposure | capillary distal | 373 | 78 | 451 | 2.784 |
| High exposure | wwp brisk | 387 | 78 | 465 | 2.747 |
| High exposure | cord stimulator | 109 | 21 | 130 | 2.730 |
| High exposure | intramedullary nail | 159 | 30 | 189 | 2.698 |
| High exposure | tender arm | 180 | 32 | 212 | 2.637 |
| High exposure | digital nerve | 132 | 23 | 155 | 2.627 |
| High exposure | brisk capillary | 443 | 78 | 521 | 2.613 |
| High exposure | prom shoulder | 157 | 27 | 184 | 2.609 |
| High exposure | neurogenic claudication | 97 | 16 | 113 | 2.587 |
| High exposure | painless rom | 104 | 17 | 121 | 2.575 |
| High exposure | day prescribed | 111 | 18 | 129 | 2.565 |
| High exposure | shaft fracture | 255 | 42 | 297 | 2.555 |
| High exposure | mandibular fracture | 175 | 28 | 203 | 2.536 |
| High exposure | epidural abscess | 329 | 53 | 382 | 2.529 |
| High exposure | tibial plateau | 1,203 | 193 | 1,396 | 2.513 |
| High exposure | plateau fracture | 757 | 120 | 877 | 2.504 |
| High exposure | anterior lateral | 137 | 21 | 158 | 2.503 |
| High exposure | wrist digits | 220 | 34 | 254 | 2.496 |
| High exposure | left tibial | 392 | 61 | 453 | 2.492 |
| Low exposure | arthralgias myalgias | 6,098 | 0 | 6,098 | -4.377 |
| Low exposure | systems denies | 5,418 | 0 | 5,418 | -4.259 |

| | | | | | |
|---|---|---|---|---|---|
| Low exposure | admission vitals | 5,387 | 0 | 5,387 | -4.253 |
| Low exposure | rdw neuts | 5,322 | 0 | 5,322 | -4.241 |
| Low exposure | breath chest | 5,278 | 0 | 5,278 | -4.233 |
| Low exposure | ast sgot | 4,736 | 0 | 4,736 | -4.124 |
| Low exposure | sgpt ast | 4,735 | 0 | 4,735 | -4.124 |
| Low exposure | habits dysuria | 4,441 | 0 | 4,441 | -4.060 |
| Low exposure | recent fevers | 4,401 | 0 | 4,401 | -4.051 |
| Low exposure | tot bili | 4,400 | 0 | 4,400 | -4.051 |
| Low exposure | procedure cardiac | 4,179 | 0 | 4,179 | -3.999 |
| Low exposure | denies prior | 4,096 | 0 | 4,096 | -3.979 |
| Low exposure | rales ronchi | 4,093 | 0 | 4,093 | -3.978 |
| Low exposure | ros denies | 4,087 | 0 | 4,087 | -3.977 |
| Low exposure | brbpr melena | 4,033 | 0 | 4,033 | -3.964 |
| Low exposure | chills rigors | 4,029 | 0 | 4,029 | -3.963 |
| Low exposure | headache vision | 3,981 | 0 | 3,981 | -3.951 |
| Low exposure | basos plt | 3,935 | 0 | 3,935 | -3.939 |
| Low exposure | headache sinus | 3,889 | 0 | 3,889 | -3.927 |
| Low exposure | nocturnal dyspnea | 3,787 | 0 | 3,787 | -3.901 |
| Low exposure | paroxysmal nocturnal | 3,778 | 0 | 3,778 | -3.898 |
| Low exposure | joint pains | 3,761 | 0 | 3,761 | -3.894 |
| Low exposure | loss vision | 3,619 | 0 | 3,619 | -3.855 |
| Low exposure | vitals transfer | 3,587 | 0 | 3,587 | -3.846 |
| Low exposure | phos tot | 3,578 | 0 | 3,578 | -3.844 |
| Low exposure | congestion sore | 3,519 | 0 | 3,519 | -3.827 |

| | | | | | |
|---|---|---|---|---|---|
| Low exposure | systems notable | 3,438 | 0 | 3,438 | -3.804 |
| Low exposure | complaint dyspnea | 3,417 | 0 | 3,417 | -3.798 |
| Low exposure | cardiac review | 3,377 | 0 | 3,377 | -3.786 |
| Low exposure | stools denies | 3,375 | 0 | 3,375 | -3.785 |

[1]Log odds represent association strength between bigram presence and extreme opioid exposure. Positive values indicate association with high exposure; negative values indicate association with low exposure.

### S4: Subgroup model performance (test set)

| Model | Subgroup | ROC-AUC | PR-AUC | Brier | Sensitivity | Specificity | PPV | NPV | Cal intercept | Cal slope |
|---|---|---|---|---|---|---|---|---|---|---|
| Ridge logistic | Surgical | 0.905 (0.892-0.918) | 0.208 (0.177-0.245) | 0.0257 (0.0233-0.0282) | 0.885 (0.847-0.949) | 0.794 (0.735-0.838) | 0.116 (0.094-0.140) | 0.996 (0.994-0.998) | 0.401 | 1.176 |
| Ridge logistic | Non-surgical | 0.902 (0.878-0.926) | 0.087 (0.057-0.129) | 0.0083 (0.0068-0.0098) | 0.819 (0.769-0.975) | 0.829 (0.642-0.880) | 0.040 (0.022-0.056) | 0.998 (0.998-1.000) | 0.874 | 1.279 |
| Random forest | Surgical | 0.908 (0.897-0.919) | 0.232 (0.196-0.275) | 0.0251 (0.0228-0.0275) | 0.888 (0.837-0.956) | 0.782 (0.712-0.825) | 0.110 (0.088-0.136) | 0.996 (0.994-0.998) | 0.184 | 1.149 |
| Random forest | Non-surgical | 0.907 (0.878-0.932) | 0.129 (0.075-0.196) | 0.0081 (0.0066-0.0096) | 0.872 (0.802-0.947) | 0.818 (0.789-0.862) | 0.040 (0.031-0.055) | 0.999 (0.998-0.999) | -0.014 | 1.044 |
| XGBoost | Surgical | 0.919 (0.910-0.929) | 0.242 (0.206-0.285) | 0.0247 (0.0225-0.0271) | 0.941 (0.905-0.975) | 0.769 (0.734-0.819) | 0.110 (0.096-0.127) | 0.998 (0.996-0.999) | 0.067 | 1.042 |
| XGBoost | Non-surgical | 0.925 (0.9 | 0.161 (0.0 | 0.0079 (0.0 | 0.851 (0.789- | 0.836 (0.726- | 0.044 (0.0 | 0.998 (0.9 | 0.412 | 1.143 |

| | | | | | | | | | | |
|---|---|---|---|---|---|---|---|---|---|---|
| | | 04-0.944) | 99-0.237) | 065-0.0093) | 0.979) | 0.901) | 26-0.069) | 98-1.000) | | |
| Notes-only | Surgical | 0.861 (0.838-0.884) | 0.191 (0.155-0.239) | 0.0253 (0.0223-0.0282) | 0.902 (0.645-0.939) | 0.645 (0.625-0.904) | 0.070 (0.064-0.175) | 0.995 (0.988-0.997) | 0.082 | 1.010 |
| Notes-only | Non-surgical | 0.812 (0.754-0.865) | 0.084 (0.033-0.165) | 0.0095 (0.0072-0.0119) | 0.865 (0.750-0.963) | 0.674 (0.603-0.773) | 0.026 (0.018-0.036) | 0.998 (0.997-0.999) | -0.794 | 0.842 |
| Combined | Surgical | 0.934 (0.920-0.946) | 0.316 (0.265-0.379) | 0.0227 (0.0201-0.0253) | 0.902 (0.855-0.946) | 0.848 (0.805-0.882) | 0.150 (0.121-0.187) | 0.997 (0.995-0.998) | 0.113 | 1.056 |
| Combined | Non-surgical | 0.908 (0.878-0.933) | 0.099 (0.052-0.193) | 0.0094 (0.0071-0.0118) | 0.923 (0.839-0.984) | 0.811 (0.802-0.865) | 0.046 (0.035-0.064) | 0.999 (0.998-1.000) | -0.365 | 0.904 |
